# Frontline-worker infections as a marker of infection-prevention stress during the 2026 Bundibugyo virus disease outbreak in the Democratic Republic of the Congo

**DOI:** 10.64898/2026.09.06.26362361

**Authors:** Johan G. L. Verheyden, Celestin Nzanzu Mudogo

## Abstract

**Objective:** To quantify the reported burden, temporal contribution, mortality and health-zone distribution of frontline-worker infections during the 2026 Bundibugyo virus disease (BVD) outbreak in the Democratic Republic of the Congo, and to examine contemporaneous infection-prevention-and-control (IPC) evidence.

**Methods:** We conducted a retrospective longitudinal analysis of DRC situation reports, the INRB-UMIE BDBV2026-Data repository, and WHO outbreak updates, per STROBE principles. National health-worker snapshots were reconstructed for 10 June-9 August 2026; detailed Ituri Province tables were analysed for 13 July-4 August. Crude case fatality among frontline and other confirmed cases was compared using risk ratios and a stratified Mantel-Haenszel odds ratio.

**Findings:** Nationally, health-worker infections rose from 16/676 confirmed cases (2.4%) on 10 June to 151/3,605 (4.2%) on 30 July, contributing 11.0% of newly accumulated cases in the first three weeks but only 2.2-2.6% thereafter. Reported case fatality was lower among health workers, nationally on 30 July (29.1% versus 44.7%; risk ratio 0.65, 95% CI 0.51-0.84) and in Ituri on 4 August (30.7% versus 42.8%; RR 0.72, 95% CI 0.56-0.92; stratified odds ratio 0.59, 95% CI 0.41-0.85). Burden was spatially concentrated but heterogeneous across health zones. IPC reports documented low facility scorecards, recurrent high-risk exposures, and inconsistent worker denominators.

**Conclusion:** Frontline-worker infections formed a substantial, front-loaded epidemic component and a visible IPC-stress marker, but aggregate data cannot establish occupational incidence or place of acquisition. Lower reported mortality likely reflects earlier recognition or care access rather than lower severity. Standardized worker denominators and linked occupational surveillance should be incorporated into Ebola situation reporting.

**What was already known on this topic:** Previous Ebola and Marburg outbreaks have demonstrated substantial health-worker involvement, with infections linked to inadequate triage, insufficient personal protective equipment, exposure to patients with unrecognized infection and fragile health systems. Occupational burden is often concentrated early in epidemics, and infections among health workers can disable essential services, erode workforce confidence and signal weaknesses in infection prevention and control. However, routine outbreak reports rarely document health-worker trajectories in detail or link them systematically to IPC performance.

**What this paper adds:** Using reconciled longitudinal national and provincial snapshots and detailed personnel de premiere ligne (PPL) tables from Ituri Province, this paper quantifies the reported burden, mortality and health-zone distribution of frontline-worker infections during the 2026 BVD outbreak. It shows that frontline infections formed a substantial, front-loaded component of the epidemic and remained a visible marker of IPC stress, while crude case-fatality ratios among frontline workers were lower than among other confirmed cases. It demonstrates that available aggregate data cannot establish occupational incidence or place of acquisition, and argues that standardized worker denominators and linked occupational surveillance should be incorporated into Ebola situation reporting.

## Introduction

Health and care workers occupy a distinctive position in Ebola epidemics. They are exposed to undifferentiated febrile illness before diagnosis, invasive clinical procedures, contaminated environments, deceased patients, and repeated contact with communities in which transmission is active. At the same time, infection among health workers can disable essential services, erode workforce confidence, and signal weaknesses in triage, personal protective equipment (PPE), environmental decontamination, or recognition of unrecognized Ebola cases. A systematic review of Ebola and Marburg outbreaks identified inadequate PPE and exposure to patients with unrecognized infection among the most frequently reported risk situations [1]. During the 2014-2016 West African epidemic, large numbers of health workers were infected even though many exposures occurred outside dedicated Ebola treatment units [2].

Bundibugyo virus disease (BVD) has its own history of occupational exposure. During the original 2007-2008 Bundibugyo outbreak in Uganda, 14 health workers were infected before strict isolation procedures were established [3]. In the 2012 BVD outbreak in the DRC, retrospective investigation identified substantial health-worker involvement, including 18 health workers among probable cases reported by mid-September [4].

The 2026 DRC outbreak produced an occupational burden on a different scale. WHO reported 16 confirmed health- and care-worker infections by 10 June, 102 by 1 July, 119 by 15 July, and 151 with 44 deaths by 30 July [5–8]. By 9 August, WHO reported at least 155 health-worker cases and 45 deaths [9]. WHO explicitly linked these infections to ongoing occupational exposure risks and persistent challenges implementing IPC in health-care facilities, particularly outside designated Ebola treatment centres [8,9].

The national totals do not answer several operational questions. Was the occupational burden concentrated early in epidemic growth or did it expand proportionately with community transmission? How were frontline infections distributed within the most affected province and health zones? How did reported mortality among frontline workers compare with other confirmed cases? Do contemporaneous IPC assessments support interpreting the frontline-worker burden as a marker of IPC stress, while avoiding the stronger and unsupported inference that every infection was acquired occupationally?

We addressed these questions using a reconciled longitudinal library of DRC situation reports and public WHO updates. We deliberately do not estimate occupational incidence or relative infection risk because stable workforce denominators are unavailable. Instead, we quantify health-worker/PPL burden as a proportion of confirmed cases, compare crude reported outcomes, describe spatial concentration, and place the epidemiological findings alongside contemporaneous IPC scorecards and exposure assessments.

## Methods

We conducted a retrospective longitudinal analysis of aggregate public outbreak surveillance data, reported in accordance with STROBE principles [10]. Source material consisted of Institut National de Sante Publique (INSP) situation reports, the INRB-UMIE BDBV2026-Data repository, and WHO Disease Outbreak News updates. The source hierarchy for DRC SitRep variables was: original INSP PDF, then INRB-UMIE manual transcription, then locally extracted SitRep text. Missing or not-determined values were never treated as zero. This secondary analysis of aggregate, non-identifiable surveillance data did not require ethics committee approval; results are not disaggregated by age or sex because source reports do not report health-worker infections at that level of detail (a limitation discussed below).

Terminology differs by source. DRC SitReps use personnels de premiere ligne (PPL), defined as health personnel and frontline service providers. WHO uses health workers or health and care workers. We retain source terminology in data tables and use ‘frontline workers’ as an umbrella term in interpretation; the categories overlap but are not assumed identical.

National snapshots were reconstructed for 10 June, 1 July, 15 July, 30 July, and 9 August. Confirmed totals and health-worker counts for the first four dates came from contemporaneous WHO updates. The 9 August health-worker count was WHO’s ‘at least 155’ estimate; the national confirmed-case denominator for that date came from the INRB-UMIE transcription of the INSP national banner and is used descriptively rather than in the primary mortality comparison. For each snapshot we calculated the health-worker share of cumulative confirmed cases, and, for successive snapshots, the frontline share of newly accumulated reported cases over the interval (not symptom-onset incidence).

Ituri was the only province with repeated health-zone PPL tables of sufficient detail for longitudinal analysis. We extracted detailed tables from SitReps covering 13 July-4 August; earlier province-level aggregate totals from 1-10 July were retained for context. SitRep 057, an isolated discontinuity (58 cumulative PPL between values of 112 and 114 on adjacent reports), was excluded. Downward reclassification occurred in early August, when the Ituri total fell from 142 to 140; we retained the reported current total for prevalence-type summaries and, for interval accumulation only, used the running maximum to avoid interpreting a downward correction as negative incident infection.

The latest detailed Ituri PPL table (4 August) was matched to the validated cumulative confirmed case/death table for the same date, giving the PPL share of confirmed cases, crude PPL case-fatality ratio (CFR), and the corresponding non-PPL CFR within each affected health zone. At national level (30 July) and in Ituri (4 August), crude reported death risk among frontline workers versus other confirmed cases was compared using risk ratios; the Ituri comparison was also stratified across the 12 PPL-affected health zones using a Mantel-Haenszel common odds ratio. These are comparisons of crude reported CFR, not patient-level causal effects.

We reviewed SitRep IPC sections for facility scorecard results, PPE/IPC supply constraints, PPL exposure-risk assessments, and PPL denominators used by the IPC pillar. Because facilities were selected non-randomly and instruments/coverage changed over time, these indicators are presented as structured operational context rather than a pooled causal IPC score. The data do not record where each PPL infection was acquired; we therefore do not classify PPL infections as nosocomial unless a source explicitly documents a health-care exposure. We did not fit lead-lag ‘sentinel’ models, because the PPL series is cumulative, irregularly updated, health-zone-detailed only after mid-July, and frontline cases are themselves embedded in the totals they might be used to predict. Analyses were performed in Python (pandas, NumPy, SciPy, statsmodels, Matplotlib).

## Results

### A front-loaded national occupational burden

The reported health-worker burden rose sharply during early expansion. On 10 June, 16 of 676 confirmed DRC cases (2.4%) were health workers; by 1 July the cumulative count had reached 102 of 1,460 (7.0%); it then eased to 119 of 2,124 (5.6%) on 15 July and 151 of 3,605 (4.2%) on 30 July (Table 1; Fig. 1). WHO reported at least 155 health-worker cases by 9 August [5–9]. The interval contribution shows how front-loaded this burden was: health workers accounted for 86 of 784 newly accumulated confirmed cases (11.0%) between 10 June and 1 July, but only 2.6% of the 1-15 July increment and 2.2% of the 15-30 July increment. Cumulative health-worker infections continued to rise, but far more slowly than the overall epidemic after early July.

**Table 1.** National health/frontline-worker burden at selected outbreak snapshots. *9 August health-worker count is WHO’s explicit lower bound (‘at least 155’); used descriptively only. Sources: WHO Disease Outbreak News DON607, DON612, DON613, DON614, DON615 [5–9].

| Date | All confirmed | Frontline confirmed | Frontline share | Frontline deaths | Frontline CFR |
| --- | --- | --- | --- | --- | --- |
| 10 Jun | 676 | 16 | 2.4% | – | – |
| 01 Jul | 1460 | 102 | 7.0% | 25 | 24.5% |
| 15 Jul | 2124 | 119 | 5.6% | 36 | 30.3% |
| 30 Jul | 3605 | 151 | 4.2% | 44 | 29.1% |
| 09 Aug* | 4381 | 155 | 3.5% | 45 | 29.0% |

**Fig. 1.**
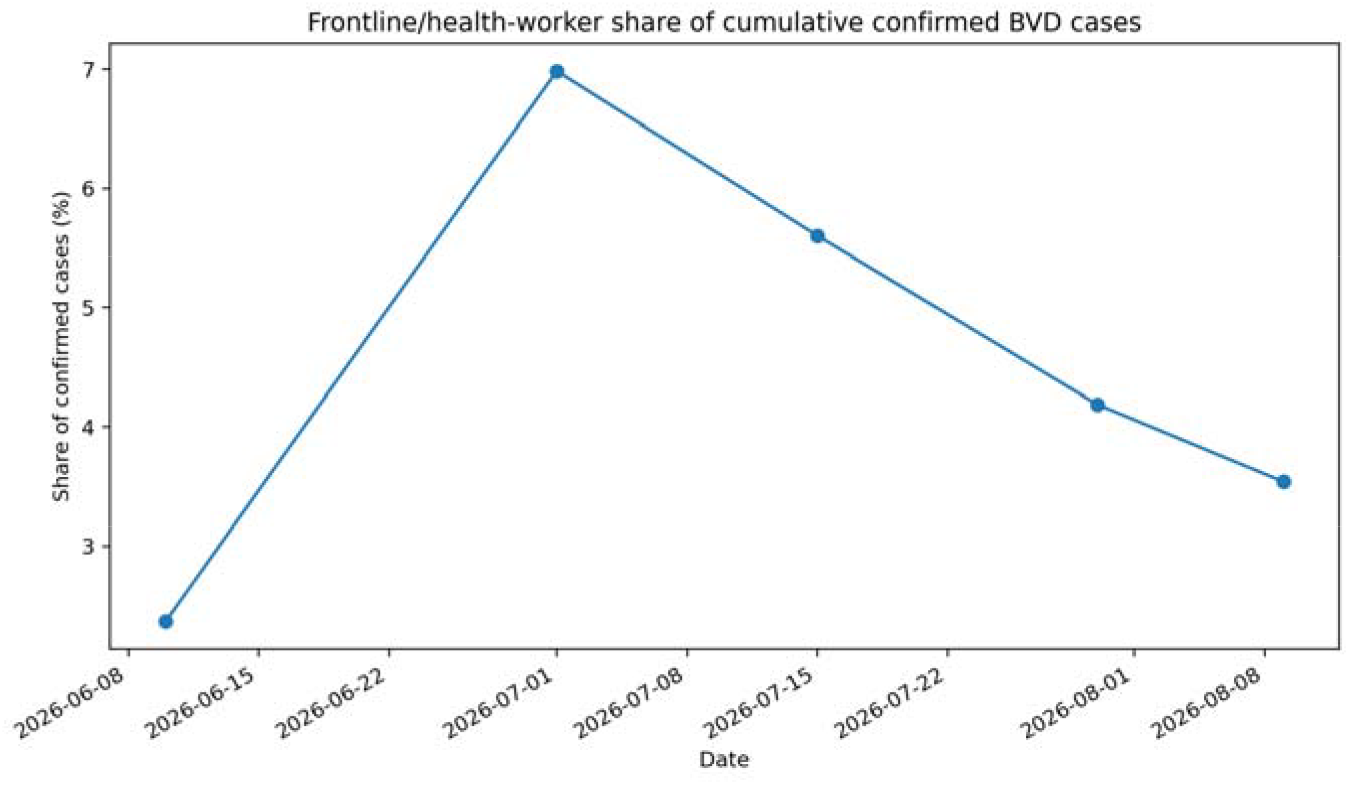
Frontline/health-worker share of cumulative confirmed BVD cases at externally validated national snapshots. The 9 August WHO count is an explicit lower bound.

### Reported frontline-worker mortality was lower than among other confirmed cases

On 30 July, 44 of 151 confirmed health workers had died (crude CFR 29.1%); among the remaining 3,454 confirmed cases, 1,543 deaths had been reported (CFR 44.7%). The crude risk ratio for death among confirmed health workers versus other confirmed cases was 0.65 (95% CI 0.51-0.84). The same pattern held in the detailed Ituri analysis: on 4 August, 43 of 140 PPL had died (30.7%), compared with 1,421 deaths among 3,320 non-PPL confirmed cases (42.8%); RR 0.72 (95% CI 0.56-0.92) (Fig. 2). Stratification across the 12 PPL-affected health zones yielded a common mortality odds ratio of 0.59 (95% CI 0.41-0.85; p=0.004). These results describe reported case fatality, not biological protection. PPL may be diagnosed earlier, monitored more intensively, reach treatment sooner, differ in age or comorbidity, or have more complete ascertainment of less severe infection; the aggregate data do not permit adjustment for these factors.

**Fig. 2.**
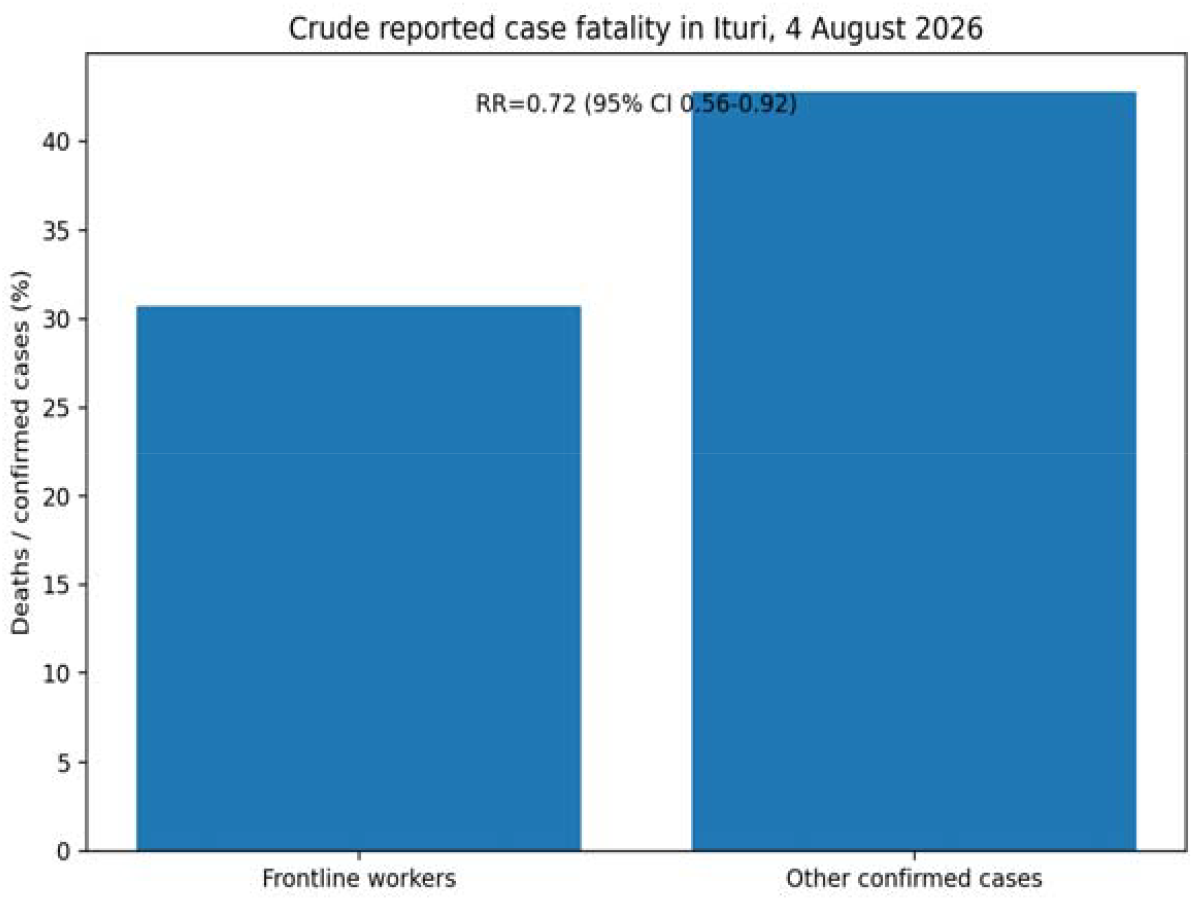
Crude reported case fatality among PPL and other confirmed cases in Ituri, 4 August 2026. The risk ratio is descriptive and unadjusted for age, timing, care access, or ascertainment.

### In Ituri, frontline cases continued to rise but became a smaller fraction of the epidemic

In Ituri, aggregate PPL counts increased from 102 on 1 July to 140 on 4 August (a 37.3% rise), while confirmed cases province-wide rose from 1,333 to 3,460 (159.6%); the PPL share of cumulative confirmed cases consequently fell from 7.7% to 4.0% (Fig. 3). The decline was more pronounced for deaths, from 6.6% to 2.9% of confirmed deaths. The detailed 13 July-4 August series nevertheless documented episodic increases, including a rise from 123 PPL on 27 July to 137 on 30 July and 141 on 31 July, before the subsequent reclassification to 140.

**Fig. 3.**
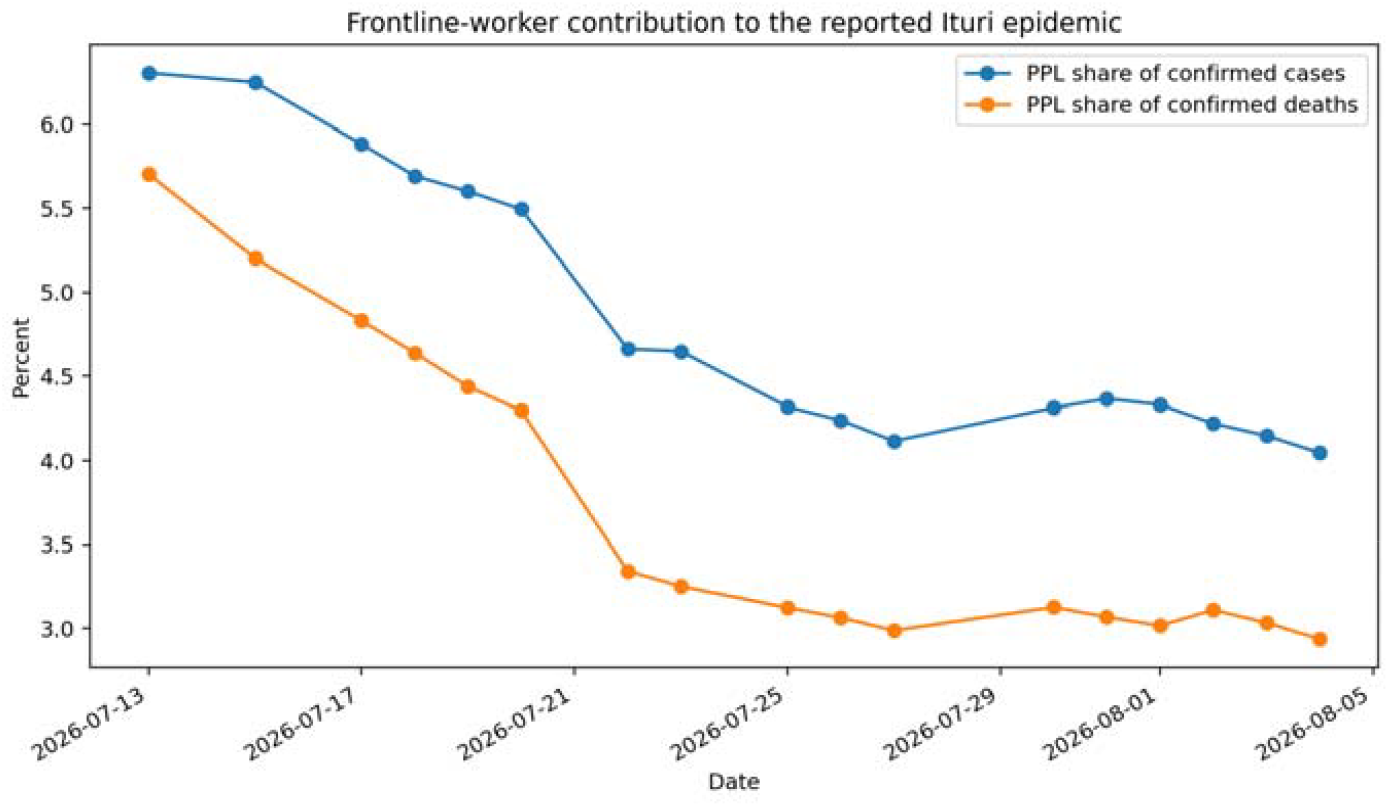
PPL contribution to cumulative confirmed cases and deaths in Ituri during the detailed PPL-table period (13 July–4 August 2026).

### Frontline-worker burden was spatially concentrated but highly heterogeneous

By 4 August, 140 Ituri PPL cases were distributed across 12 health zones. Bunia (35), Rwampara (33), Nia-Nia (22), Mongbwalu (17) and Nyankunde (14) accounted for 121/140 (86.4%) of all reported PPL cases (Table 2). However, the PPL proportion of all confirmed cases varied markedly by zone, from 0.7% in Nizi to 30.0% in Logo, 19.6% in Nia-Nia, and 12.2% in Nyankunde (Fig. 4). These proportions cannot be interpreted as occupational attack rates, since the size and composition of the frontline workforce in each zone are unknown; they do, however, show that occupationally relevant case burden was not a fixed fraction of community case burden.

**Table 2.** PPL (personnel de première ligne) burden by affected Ituri health zone, 4 August 2026. Small-denominator zones (e.g. Logo, n=10 total cases; Damas, n=18) produce unstable percentages and should be read with caution.

| Health zone | PPL cases | PPL deaths | All cases | All deaths | PPL share | PPL CFR |
| --- | --- | --- | --- | --- | --- | --- |
| Bunia | 35 | 14 | 950 | 289 | 3.7% | 40.0% |
| Rwampara | 33 | 7 | 692 | 282 | 4.8% | 21.2% |
| Nia-Nia | 22 | 5 | 112 | 66 | 19.6% | 22.7% |
| Mongbwalu | 17 | 9 | 558 | 268 | 3.0% | 52.9% |
| Nyankunde | 14 | 2 | 115 | 35 | 12.2% | 14.3% |
| Lita | 4 | 1 | 151 | 89 | 2.6% | 25.0% |
| Mangala | 4 | 1 | 108 | 67 | 3.7% | 25.0% |
| Nizi | 3 | 3 | 431 | 211 | 0.7% | 100.0% |
| Logo | 3 | 0 | 10 | 5 | 30.0% | 0.0% |
| Bambu | 2 | 0 | 69 | 17 | 2.9% | 0.0% |
| Kilo | 2 | 0 | 29 | 11 | 6.9% | 0.0% |
| Damas | 1 | 1 | 18 | 8 | 5.6% | 100.0% |

**Fig. 4.**
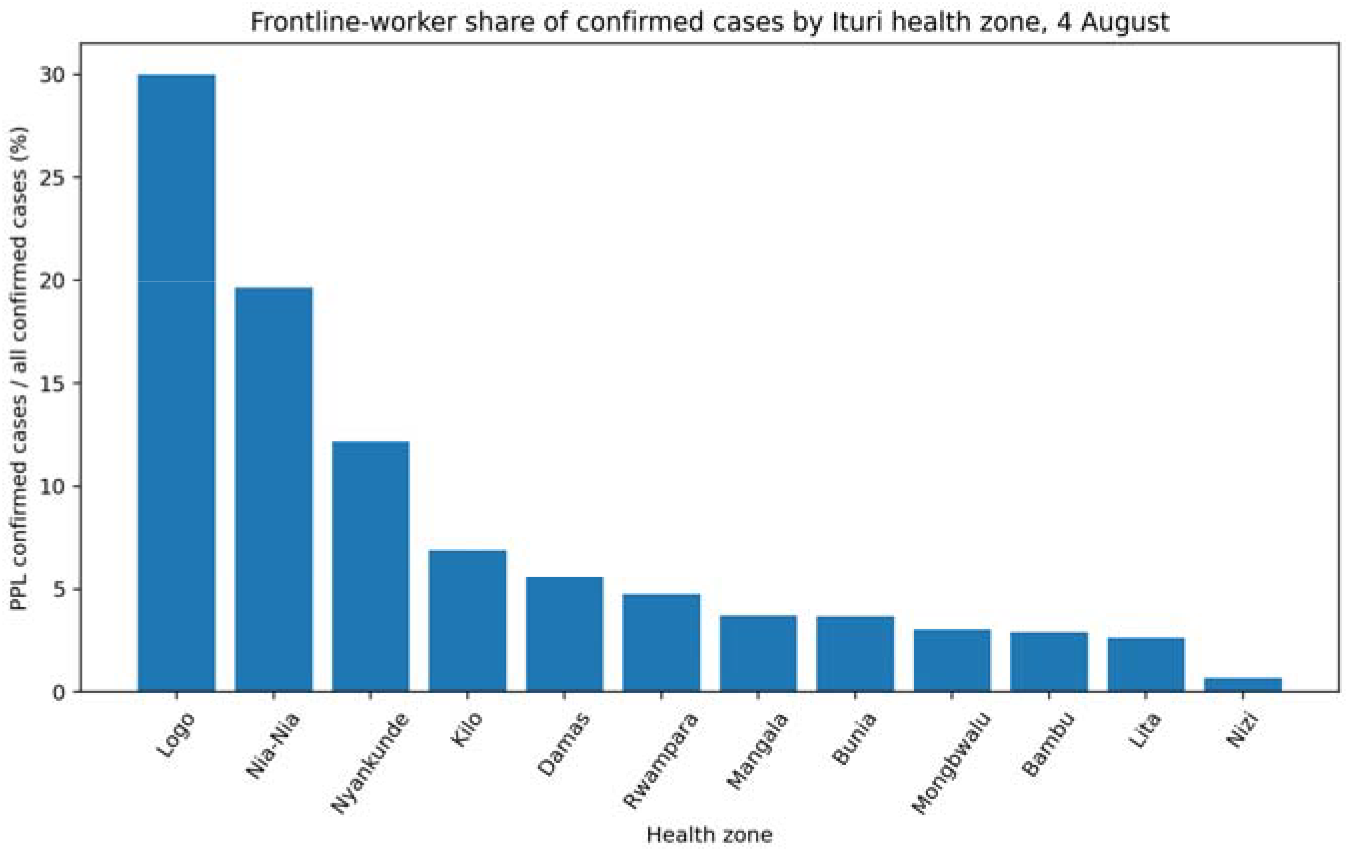
PPL share of confirmed cases across the 12 Ituri health zones reporting PPL infections on 4 August. These percentages are not occupational incidence rates.

### IPC evidence showed persistent vulnerabilities and incomplete denominator harmonization

The SitRep IPC record documents persistent vulnerability. At outbreak onset, scorecards were 7% at CS Abelkozo and 34% at HGR Mongbwalu; in Nyankunde, a facility scored 28% while 18 suspected patients were managed without isolation, including eight care providers. Later Ituri assessments continued to document low scores, including 11-12% in Mongbwalu facilities and multiple Bunia and Aru facilities below 50% (Table 3). Nord-Kivu risk-assessment data similarly documented high-risk occupational exposures: 74 of 108 assessed PPL (68.5%) were classified high-risk around 1 July, and 58 of 79 (73.4%) around 3 August; these proportions apply only to assessed workers, not population attack rates. The source systems were not fully harmonized. Around end-July, the IPC pillar reported 134 PPL contaminated among 2,884 exposed (4.6%), while the surveillance table reported 141-142 confirmed PPL in Ituri (subsequently revised to 140). Occupational denominators and case registries were being maintained by different response pillars and should not be silently combined.

**Table 3.**
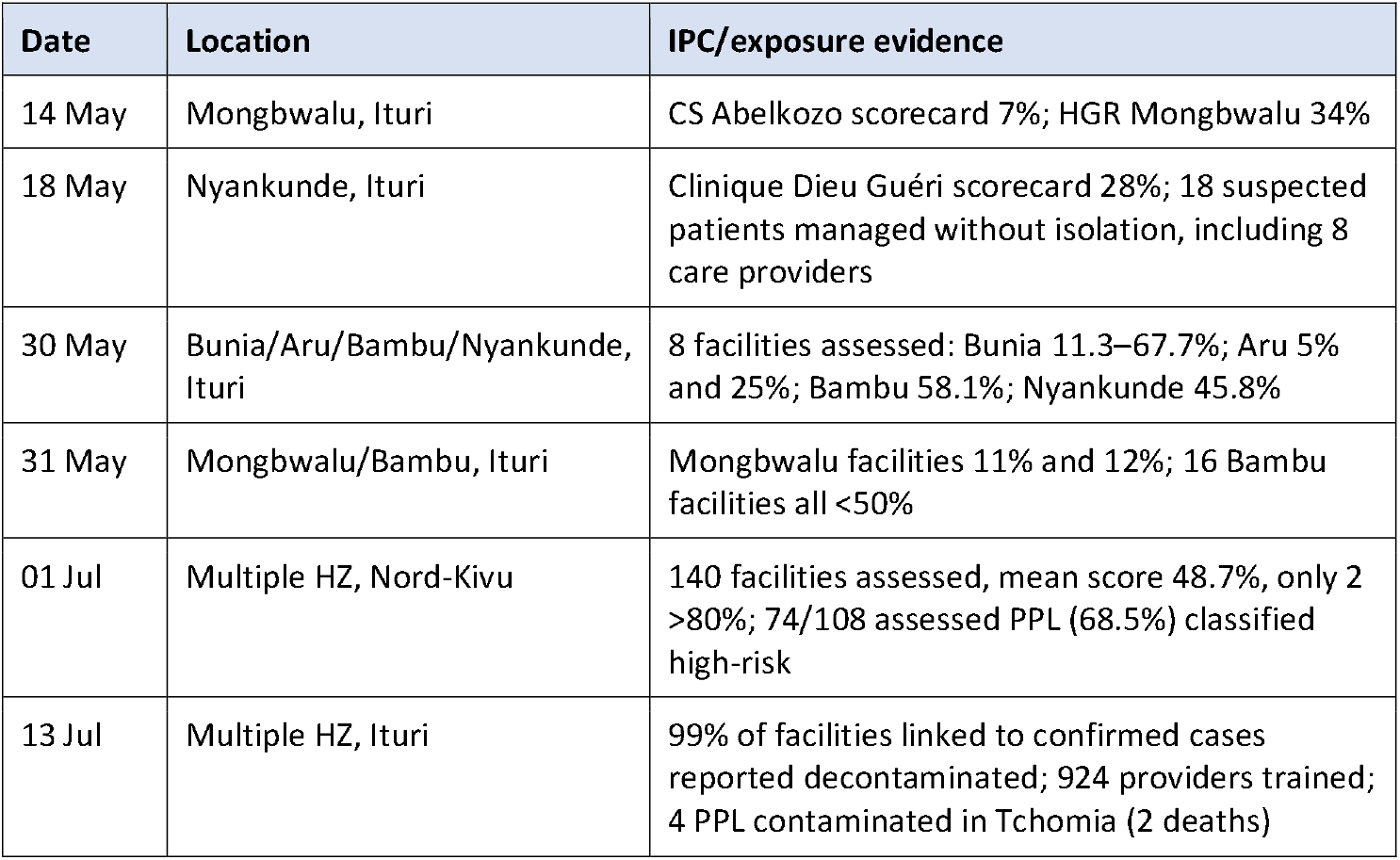
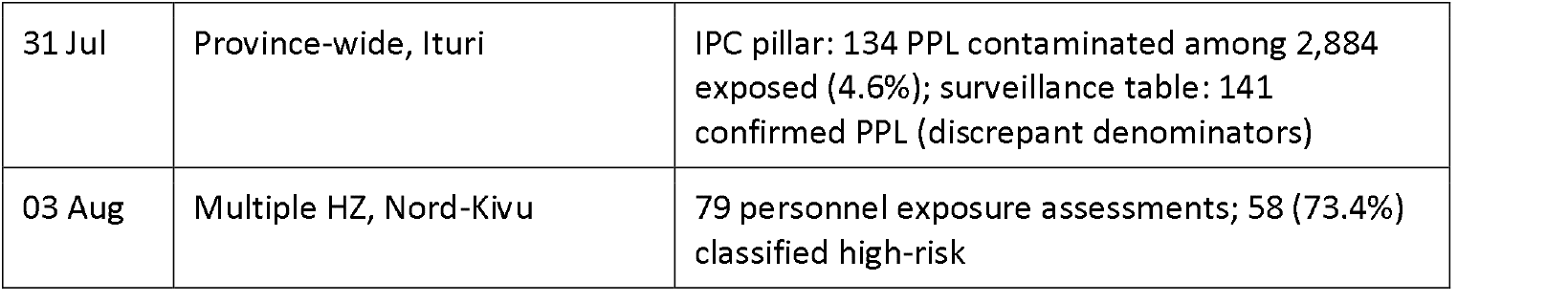
Selected infection-prevention-and-control (IPC) and occupational-exposure evidence from situation reports. Facilities were selected non-randomly and reporting coverage changed over time; figures are structured operational context, not a representative province-level IPC score.

## Discussion

This study documents a large but temporally changing frontline-worker burden. Health-worker infections rose from 16 reported cases on 10 June to more than 100 by 1 July; during those three weeks, health workers accounted for approximately one in nine newly accumulated confirmed cases. Thereafter, the absolute number continued to increase but the occupational share of the expanding epidemic fell rapidly, consistent with a front-loaded occupational shock during a period when unrecognized disease was circulating through ordinary health-care pathways while IPC systems were still being reinforced.

That interpretation is supported, but not proven, by the IPC record: early affected facilities had strikingly low scorecard results, suspected patients were sometimes managed without isolation, PPE/IPC supply gaps were repeatedly documented, and a large fraction of selectively assessed workers were classified as high-risk exposures - consistent with the established Ebola literature, in which exposure to unrecognized infected patients and inadequate PPE or infection-control infrastructure repeatedly contribute to health-worker infections [1,2]. The original 2007 Bundibugyo outbreak itself infected 14 health workers before strict isolation procedures were established [3].

The declining PPL share of reported cases should not be read as proof that occupational risk became low. Community transmission expanded much faster than the PPL numerator; IPC practice may have improved; earlier recognition and triage may have reduced exposures; the most susceptible facilities may already have experienced clusters; and PPL-reporting definition or completeness may have changed. Continued reports of high-risk exposures in Nord-Kivu and WHO’s finding that health-worker infections were still increasing in August argue against reading the declining proportion as resolution of occupational risk [8,9].

The lower reported CFR among PPL is plausible if health workers were recognized earlier, tested more readily, admitted sooner, or had better access to optimized supportive care; it may also reflect age, health status, case-ascertainment intensity, or differential outcome reporting. The aggregate SitRep data cannot distinguish these explanations, and we do not interpret the mortality ratio as a causal protective effect of frontline status or as evidence that BDBV is biologically less severe in health workers.

The spatial analysis cautions against treating health-worker infections as a fixed proportion of epidemic size: although Bunia and Rwampara held the largest absolute PPL burdens, smaller zones such as Nia-Nia, Nyankunde and Logo had much higher PPL proportions among their confirmed cases, patterns that can arise from small denominators, local facility clusters, or differences in workforce composition or ascertainment. Without workforce denominators, these cannot be converted into incidence rates, but they are useful operational signals that particular health systems may warrant targeted IPC investigation even when total case numbers are modest.

We initially considered modelling frontline infection as a leading sentinel of subsequent community transmission, but the available data do not support that estimand: detailed PPL counts begin late, are cumulative rather than onset-dated, updated irregularly, and are themselves embedded in the health-zone totals they might be used to predict. A more defensible use of PPL infections is as a marker of occupational exposure and IPC stress that should trigger investigation, not as a quantified predictor of community incidence.

The data architecture revealed here deserves attention: the surveillance and IPC pillars reported different PPL totals and denominators. The IPC pillar’s 134-contaminated-among-2,884-exposed figure is potentially valuable as one of the few attempts to provide an exposure denominator, but its case count did not reconcile with the surveillance register. Future Ebola SitReps should publish a linked occupational line list, or at minimum a stable table of worker category, facility, health zone, onset and confirmation dates, probable exposure setting, exposure-risk category, outcome, and denominator of workers at risk; WHO’s 2025 occupational-exposure guidance already provides a framework for risk categorization [11].

The study has important limitations. PPL and WHO health-worker categories may not be identical. No stable total workforce denominator is available by health zone, so occupational incidence and relative infection risk cannot be estimated. Place of acquisition is unknown for most cases, and results are not disaggregated by age or sex because source reports do not provide this detail. Cumulative counts are subject to retrospective revision and delayed reporting; outcome completeness is imperfect, and crude-CFR comparisons can be affected by differential follow-up and ascertainment. Facility scorecards were performed selectively, with changing coverage, so cannot be treated as representative province-level IPC scores. The detailed PPL health-zone series is concentrated in Ituri and ends 4 August even though national health-worker infections continued thereafter.

Despite these limitations, DRC SitRep totals and WHO’s independent updates are externally consistent at key dates: on 30 July, the DRC source decomposition of Ituri, Nord-Kivu and Haut-Uele PPL counts reconciles to WHO’s national total of 151 health-worker infections and 44 deaths, supporting the use of the detailed SitRep tables for describing occupational burden while retaining the documented data-quality caveats.

### Conclusion

Frontline-worker infections were a substantial and front-loaded component of the 2026 BVD epidemic in the DRC. Their share of reported cases fell as community transmission expanded, but occupational infections continued and were accompanied by persistent IPC vulnerabilities and high-risk exposure assessments. Reported frontline-worker CFR was lower than among other confirmed cases, a difference consistent with earlier recognition or care access but not causally explained by aggregate data. Frontline-worker infections should be treated as an IPC warning signal requiring rapid occupational investigation, while future outbreak reporting should provide harmonized worker denominators and linked exposure data so that true occupational risk can be estimated.

## Data Availability

All data produced in the present study are available upon reasonable request to the authors

## Declarations

### Ethics approval

This study used publicly available, aggregate, non-identifiable outbreak surveillance data. No individual-level records were accessed; institutional ethics review and individual consent were therefore not required for this secondary analysis.

### Data availability

Public source data are available through the DRC INSP Ebola SitRep archive, WHO Disease Outbreak News, and the INRB-UMIE BDBV2026-Data repository. Analysis-ready tables and derived results supporting this manuscript are included in an accompanying reproducibility package, available on request.

### Author contributions

J.G.L.V.: conceptualization, methodology, data curation, formal analysis, visualization, writing — original draft. C.N.M.: epidemiological and clinical interpretation, validation, writing — review and editing. Both authors approved the final manuscript and, per ICMJE criteria, take public responsibility for its content.

### Declaration of AI assistance

During manuscript preparation, the authors used Claude (Anthropic; Claude Sonnet 5) under supervision to assist with data tabulation, organisation, language polishing and formatting. All analytic definitions, numerical results, interpretations and final text were reviewed and accepted by the authors, who take full responsibility for the accuracy and integrity of the work.

